# Gabapentinoid Use among Medicare Beneficiaries during the Post-Stroke Recovery Period

**DOI:** 10.1101/2025.08.06.25333178

**Authors:** Julianne D. Brooks, Rebeka Bustamante Rocha, Maria A. Donahue, Joseph P. Newhouse, Sebastien Haneuse, Alexander C Tsai, Lidia M.V.R. Moura

## Abstract

**Background:** Most studies of gabapentinoid prescribing trends have focused on younger, commercially insured populations. Patterns among vulnerable groups, including older stroke survivors, remain poorly characterized. We described patterns of gabapentinoid initiation among Medicare beneficiaries following acute ischemic stroke (AIS).

**Methods:** We analyzed claims data from a 20% sample of U.S. Medicare beneficiaries aged ≥ 65 years hospitalized for AIS between 2009 and 2022. We included individuals enrolled in traditional Medicare for ≥ 12 months before hospitalization, without prior stroke during this interval, and discharged home or to an inpatient destination for ≤30 days. After excluding those with gabapentinoid use before stroke, we analyzed outpatient gabapentin prescriptions six months after stroke discharge by demographic and clinical characteristics. We calculated the percentage of gabapentinoid initiators by year and U.S. census division, standardized by age, discharge destination, and modified Rankin Score (mRS).

**Results:** Among 153,728 Medicare stroke survivors who had not previously been prescribed a gabapentinoid in the 6 months before hospitalization, 4.9% received new prescriptions within 6 months after discharge. The median age was 78 years (Quartile Range: 72 - 84), 55% were female, and 81% were Non-Hispanic White. The crude percentage of gabapentinoid initiators increased from 3.6% in 2009 to 5.8% in 2022. The standardized percentages were 3.8% in 2009 and 5.9% in 2022. We reported a 2.1 percentage point difference in gabapentinoid initiation between U.S. census divisions, with the lowest percentage in the New England Division (3.9%) and the highest in the West South Central Division (6.0%). After standardization, the percentage point difference was 1.9% between the highest and lowest U.S. division.

**Conclusion:** The percentage of U.S. Medicare stroke survivors initiating gabapentinoid increased from 2009 to 2022. We also identified geographic variation, with the highest initiation percentage in the West South Central and the lowest in the New England U.S. Division.

Twitter: @MGHValue

## INTRODUCTION

Stroke is a leading cause of long-term disability. In the United States, over 795,000 people have a stroke each year, with 610,000 being new events.^1^ Chronic pain is a common and often debilitating complication of stroke,^2,3^ affecting up to 30% of stroke survivors.^4^ The risk of chronic pain is greater during the subacute and chronic phases of recovery, particularly among older adults and those with greater stroke severity.^4^ Common types of post-stroke pain include shoulder pain, central post-stroke pain (CPSP), painful spasticity, and tension-type headache, all of which are associated with a reduced quality of life.^5^

Gabapentinoids (gabapentin and pregabalin) are antiseizure medications frequently prescribed for pain management.^6^ Initially approved by the U.S. Food and Drug Administration (FDA) for the treatment of seizures, these drugs later gained approval for specific pain-related conditions. Both gabapentin and pregabalin are approved for the treatment of postherpetic neuralgia, while pregabalin is also approved for the treatment of diabetic neuropathy, fibromyalgia, and pain associated with spinal cord injury.^7,8^ Notably, gabapentinoid use among older adults remains controversial given their heightened vulnerability to adverse effects commonly associated with gabapentinoids, including delirium, cognitive impairment, dizziness, and increased risk of falls.^9,10^

Gabapentinoid prescriptions in the U.S. have surged substantially over the past two decades, rising from 1.2% of all adults in 2002 to 4.7% in 2021.^11,12^ This increase may reflect their clinical endorsement, especially in the setting of neuropathic pain, although gabapentin use in this context remains off-label.^13,14^ Despite having fewer FDA-approved indications than pregabalin, gabapentin is more frequently prescribed and was ranked the 6^th^ most commonly dispensed drug in the U.S. in 2018.^15^ One analysis of the U.S. National Ambulatory Medical Care Survey estimated that 98.3% of gabapentin prescriptions in 2003-2016 were for off-label use^16^ (i.e., to treat conditions such as anxiety, insomnia, alcohol use disorder, chronic cough, and hot flashes, for which robust, high-quality, definitive evidence supporting effectiveness is lacking or inconsistent^6^).

The high prevalence of post-stroke pain and the increasing reliance on gabapentinoids for the management of post-stroke pain raise important questions about the appropriateness of this use in vulnerable populations. While previous studies have identified demographic and regional variation in prescriptions, most have relied on data from younger and commercially insured populations.^17,18^ Patterns of use among high-risk groups, such as older stroke survivors, remain poorly characterized. Understanding geographic and temporal variation in gabapentinoid use in this population would offer insights into differences in clinical practice and quality of care. Therefore, our study aimed to characterize gabapentinoid initiation patterns among Medicare beneficiaries aged 65 years or older following acute ischemic stroke (AIS).

## METHODS

This study was approved by the Mass General Brigham Institutional Review Board and followed the Strengthening the Reporting of Observational Studies in Epidemiology (STROBE) reporting guidelines (**Supplemental Table S1**).^19^ The requirement for informed consent was waived in our study, as we conducted a secondary analysis of data routinely collected for billing purposes. The data supporting this study’s findings were collected by the U.S. Centers for Medicare & Medicaid Services (CMS) and were made available by CMS with no direct identifiers.^20^ All results were aggregated following the CMS Cell Size Suppression Policy (version dated January 26, 2024). We cannot share the data, which were used under license from CMS for this study, but our analysis code is provided in the supplemental materials (Supplementary Materials – analytical code).

### Study Design

We conducted a retrospective analysis of a 20% random sample of U.S. Medicare claims data from January 1, 2008, to December 31, 2022. We included adults aged 65 and over who were discharged home after a hospitalization for AIS between January 1, 2009, and June 30, 2022, to allow for at least 1 year of pre-stroke data and 6 months of post-stroke data.

Hospitalizations were selected from the Medicare Provider Analysis and Review (MedPAR) database based on principal diagnosis codes for AIS^21^: International Classification of Diseases, 9th Revision (ICD-9) codes 433, 434, 436, and ICD-10 code I63, a validated strategy to capture AIS in administrative databases.^22^

We included Medicare beneficiaries who were enrolled in traditional Medicare Part A (hospital insurance), Part B (medical insurance), and Part D (drug prescription coverage) continuously for at least 12 months before their admission for stroke. We included first stroke admissions, using at least one year of look back to identify prior stroke.^23^ We focused on outpatient gabapentinoid initiation among community-dwelling stroke survivors. We included individuals discharged home and those discharged to a skilled nursing facility (SNF), rehabilitation, or other inpatient facility and discharged home within 30 days of the index hospitalization discharge date. Our sampling strategy is shown in **Figure 1**.

**Figure 1.**
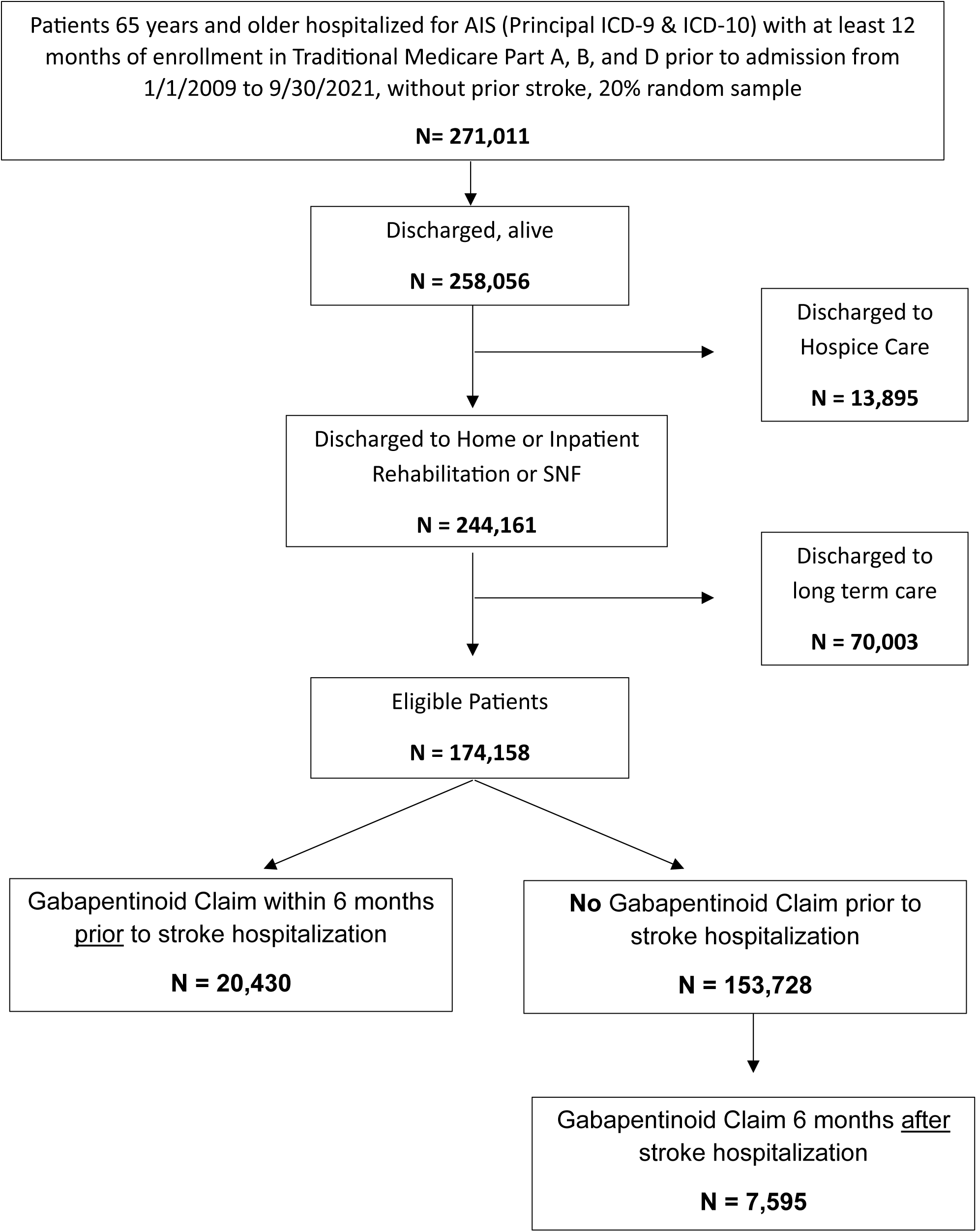
Sampling Strategy; AIS, Acute Ischemic Stroke

### Participant Characteristics

We analyzed the distribution of age, sex, race, and ethnicity in the sample. Reported race and ethnicity (Non-Hispanic White, Non-Hispanic Black, Hispanic, Asian, Other) were categorized using the Research Triangle Institute (RTI) race/ethnicity variable provided in the Master Beneficiary Summary File (MBSF).^24^ We identified baseline comorbid conditions using Medicare’s Chronic Condition Warehouse (CCW).^25^ Study characteristics are presented in **Table 1**. For this study, we used a validated claims-based algorithm to classify modified Rankin Scores (mRS) as a measure of disability. The mRS has seven total categories ranging from no or low disability to death: 0 (no symptoms), 1 (no significant disability), 2 (slight disability), 3 (moderate disability), 4 (moderate to severe disability), 5 (severe disability), 6 (death).^26,27^ We classified mRS as a binary outcome, grouping scores of 0 to 3 and 4 to 6 (i.e., ambulatory vs. non-ambulatory^28^). The algorithm could accurately identify disability status with an ROC AUC of 0.85.^29^

**Table 1.**
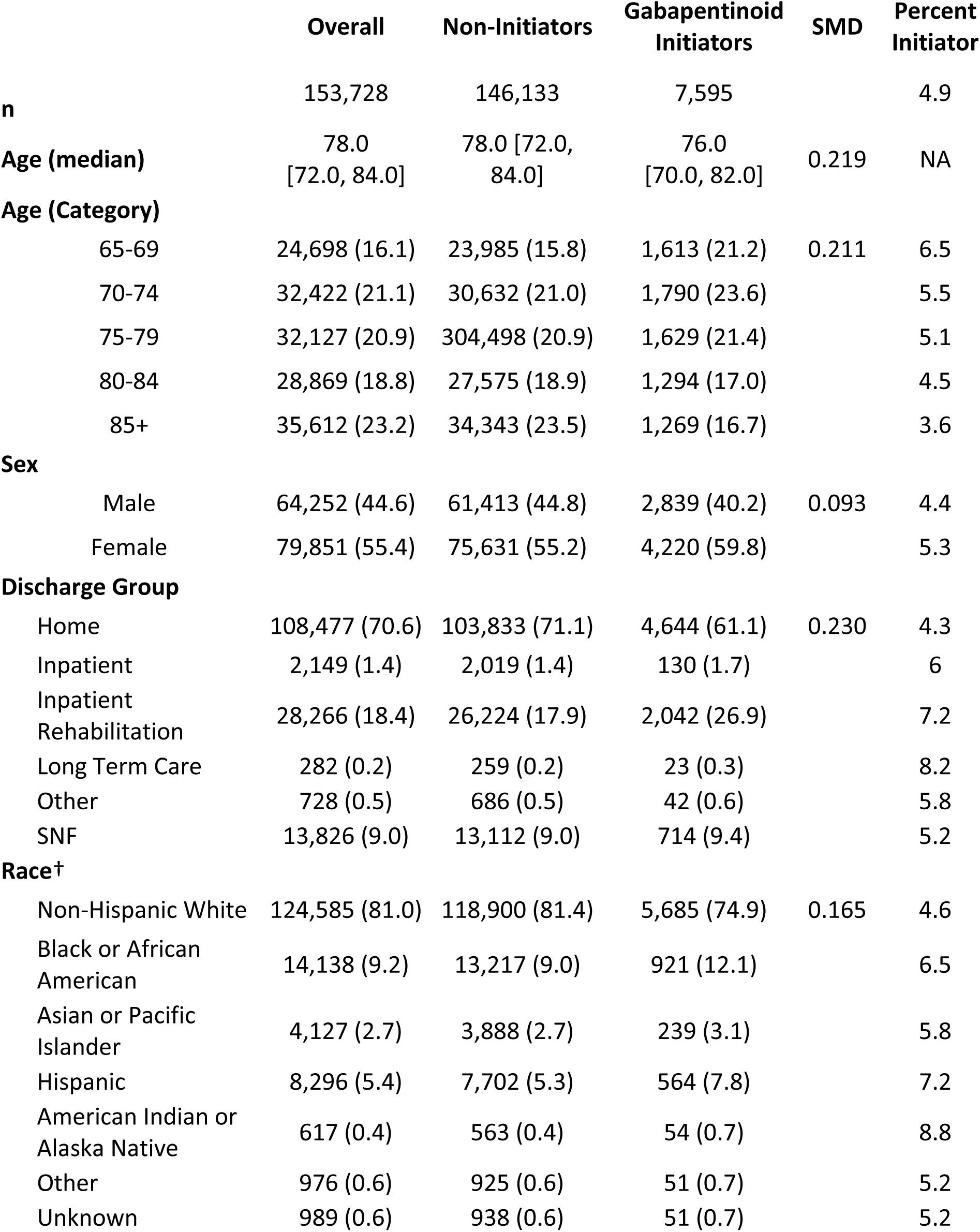
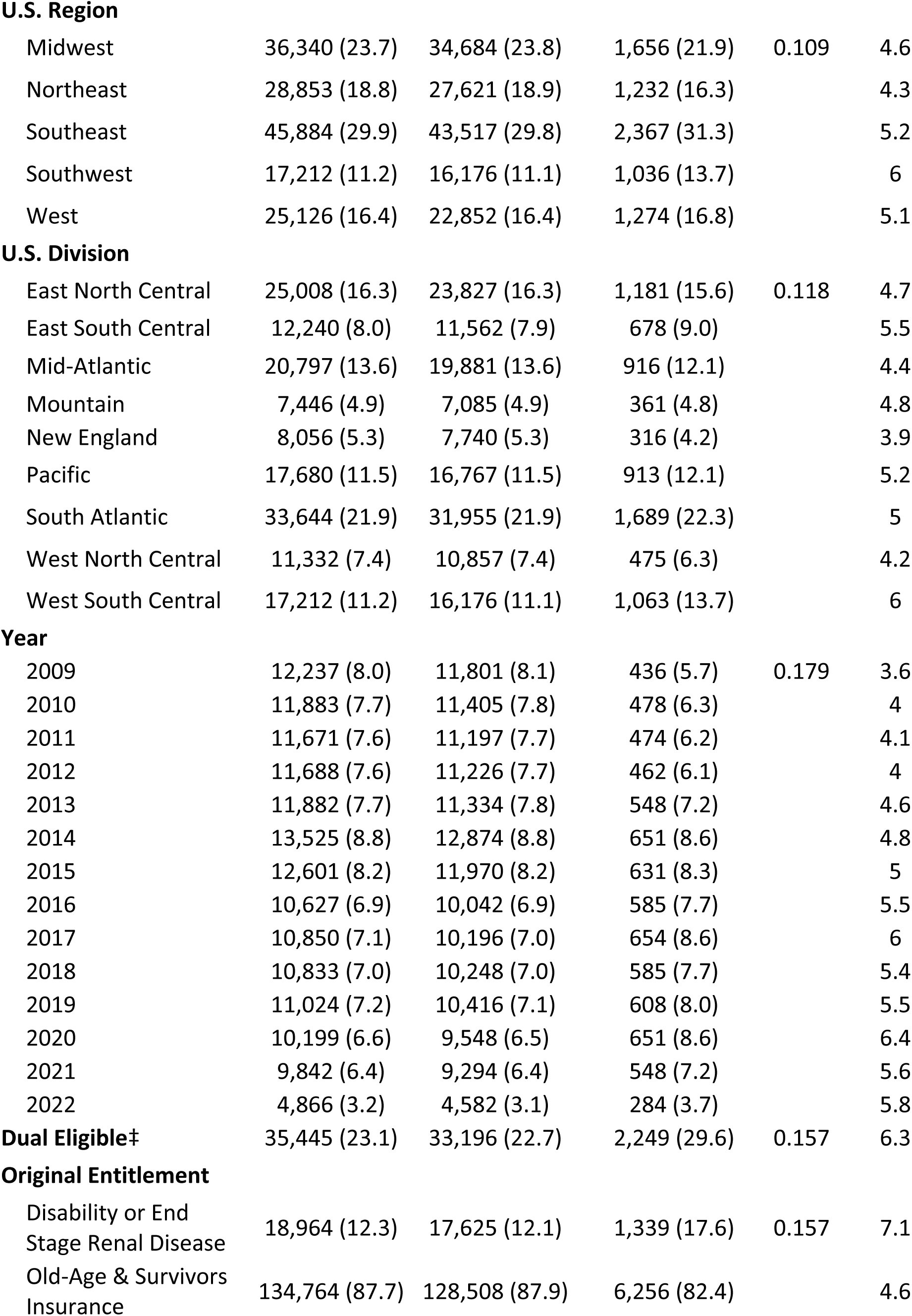
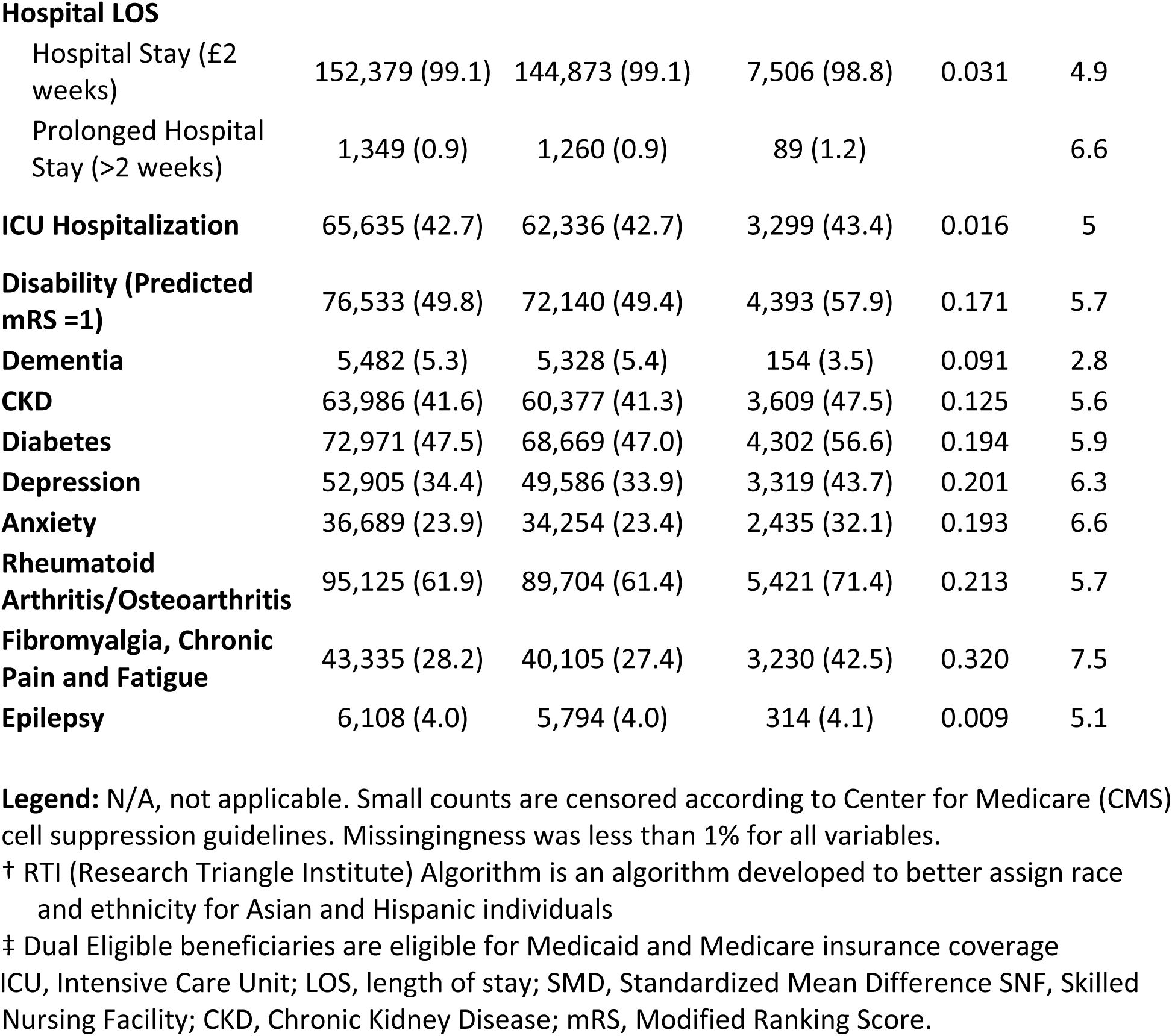
Sample Characteristics and Percent of Incident Gabapentinoid Users

### Gabapentinoid Analysis

We calculated the number of individuals with a gabapentinoid (gabapentin or pregabalin) prescription 180 days (6 months) before and after stroke hospitalization. Individuals were followed from the day discharge to the first gabapentinoid prescription claim or the end of the study observation period (180 days after discharge). Using Medicare claims data, we were able to follow individuals to the end of the study period. After removing those individuals who were previously prescribed gabapentinoids, we calculated the percentage of gabapentinoid initiators by the covariates in **Table 1**. For those who initiated gabapentinoids, we described characteristics of the prescription claims, including medication choice (gabapentin vs. pregabalin) and prescriber specialty.

We calculated adjusted gabapentinoid initiation percentages by year and US division. To that end, we first fit a logistic regression and included year, US division, age, mRS score, and discharge destination ( a, home; b, inpatient rehabilitation and then home in 30 days; or c, other inpatient location and then home in 30 days) as model covariates. We used a cubic b-spline term for year with four knots. Second, we estimated year-specific percentages by averaging predictions from the fitted model, but with the covariate distribution in each year set to the observed covariate distribution in 2021; we standardized to the covariate distribution in 2021. We repeated the process for U.S. Divisions, calculating the predicted probabilities for each U.S. division, standardizing to the covariate distribution for the East North Central Division.

## RESULTS

Among 174,158 U.S. Medicare beneficiaries discharged home within 30 days after hospitalization for AIS, we excluded 20,430 who had received a gabapentinoid prescription in the 6 months before hospitalization (**Figure 1**). In the analytic sample of 153,728 Medicare stroke survivors without prior gabapentinoid use, 4.9% received new prescriptions within 180 days after discharge. Sample characteristics are shown in **Table 1**. The median age was 78 years (QR:72-84), 55% were female, and 81% were Non-Hispanic White. A total of 23% of the sample was dually eligible for Medicaid, and most (88%) were eligible for Medicare due to age or survivors’ insurance. Nearly all of their hospitalizations (99%) had a length of stay no greater than two weeks, and 43% had an intensive care unit (ICU) stay during hospitalization. We reported the percentage of new gabapentinoid initiators in **Table 1**. The frequency of chronic conditions present at baseline is reported in **Table 1** and **Supplemental Table S2**.

### Drug Choice and Prescriber Types

Gabapentin was prescribed more often than pregabalin; 90% of new initiators received gabapentin and 10% received pregabalin prescriptions. Of the new gabapentinoid initiators (N = 7,595), the specialty of the initial prescriber was most frequently medicine (77%) and neurology (13%) (**Table 2**).

**Table 2.** Prescribers for First Prescription

| Prescriber Type | Count (%) |
| --- | --- |
| Medicine | 5,414 (76.7) |
| Neurology | 937 (13.3) |
| Psychiatry | 34 (0.5) |
| Other | 661 (9.4) |
| Missing | 549 (7.8) |

### Initiation Patterns by Clinical Characteristics

Following AIS hospitalization, most patients (71%) were discharged directly home, while 18% were discharged to inpatient rehabilitation, and 11% were discharged to other inpatient settings and then discharged home within 30 days of the index hospitalization. The percentage of gabapentinoid initiation varied by discharge destination: home (4.3%), inpatient hospital (6.0%), inpatient rehabilitation (7.2%), long-term care (8.2%), skilled nursing facility (5.2%), and other (5.8%) (SMD: 0.23). As shown in Table 1, the gabapentinoid initiation percentage decreased with advancing age. Individuals aged 65-69 years had a 6.5% initiation percentage, while individuals aged 85+ years had a 3.6% initiation rate (SMD: 0.21). Using a 0.2 threshold for defining a substantial difference based on standardized mean differences^30^, there were 3 conditions in which the gabapentinoid initiation percentage differed substantially among those with vs. without the condition: depression, fibromyalgia/chronic pain/fatigue, and rheumatoid arthritis/osteoarthritis (**Table 1**). Other baseline comorbidities are shown in the **Supplemental Materials Table S2.**

### Initiation Trend

The percentage of gabapentinoid initiation increased from 3.6% in 2009 to 5.8% in 2022. The highest percentage (6.4%) was observed in 2020 (**Table 1**, **Figure 2**). After standardization, we observed a monotonically increasing percentage over time (**Figure 2**). We presented the percentage of the sample with indication for Gabapentinoids at baseline by year and additional covariates by year in the **Supplemental Materials, Figure S1, and Table S3.** In the overall sample, the percentage of individuals with baseline anxiety and fibromyalgia, chronic pain, and fatigue increased over time.

**Figure 2.**
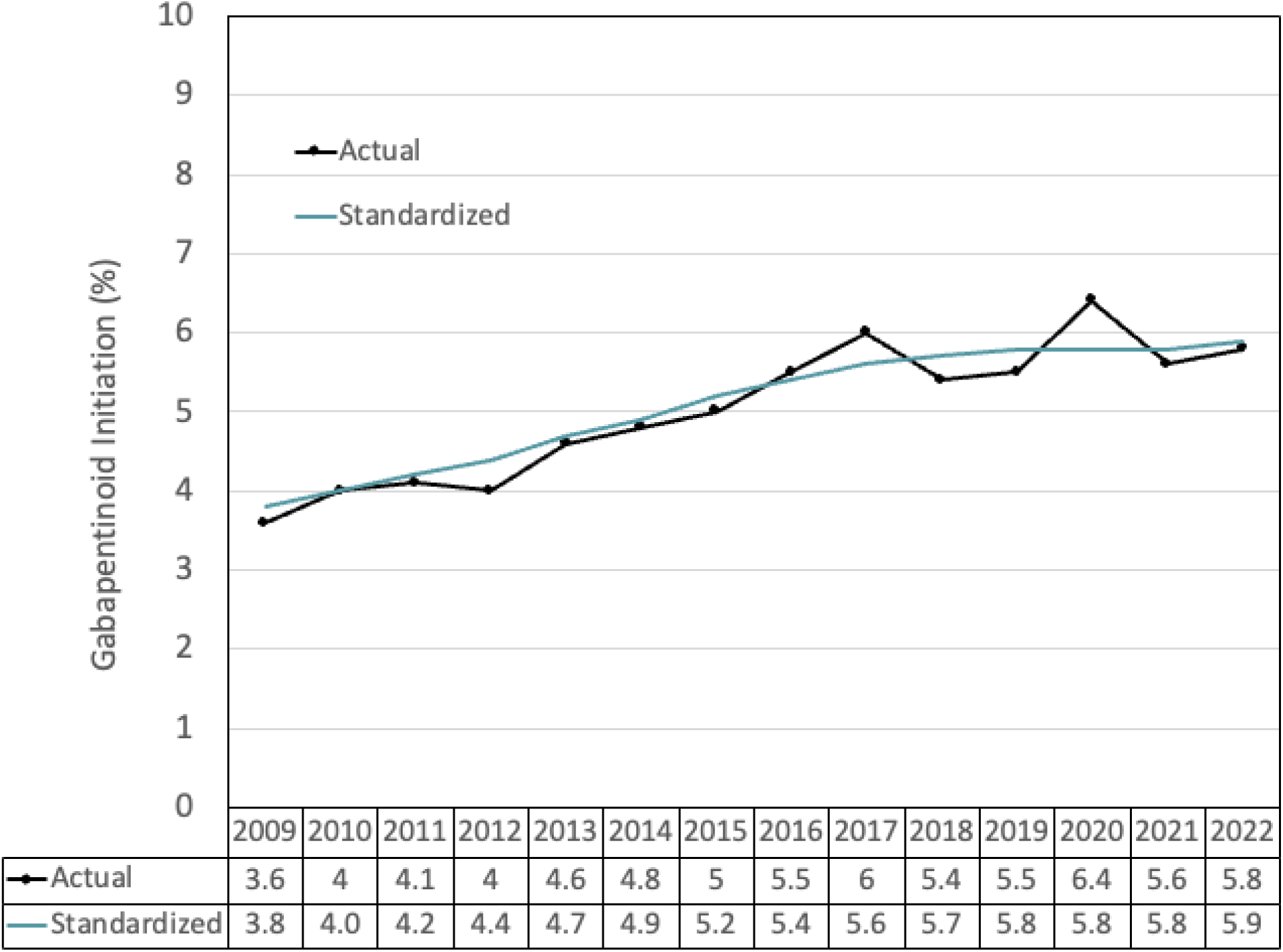
Percentage of Incident Gabapentinoid Initiators within 180 days after Stroke Discharge by Year. Standardized Percentage was standardized by Age, US Division, mRS, and Discharge Destination using 2021 covariate distribution

### Initiation Patterns by Geography

Variation by U.S. census division was observed, with the highest crude gabapentinoid percent initiation in the West South Central division (6.0%) and the lowest in New England (3.9%). After standardizing the percentages to the East North Central covariate distribution for age, disability status, discharge destination, and year trends, the highest and lowest point estimates for the percent of gabapentinoid initiators by region were 5.9% for West South Central and 4.0% for New England (Figure 3). We showed the distribution of select covariates by U.S. Division in **Supplemental Table S4,** and the model outputs used to standardize percentages are shown in **Supplemental Table S5.**

**Figure 3.**
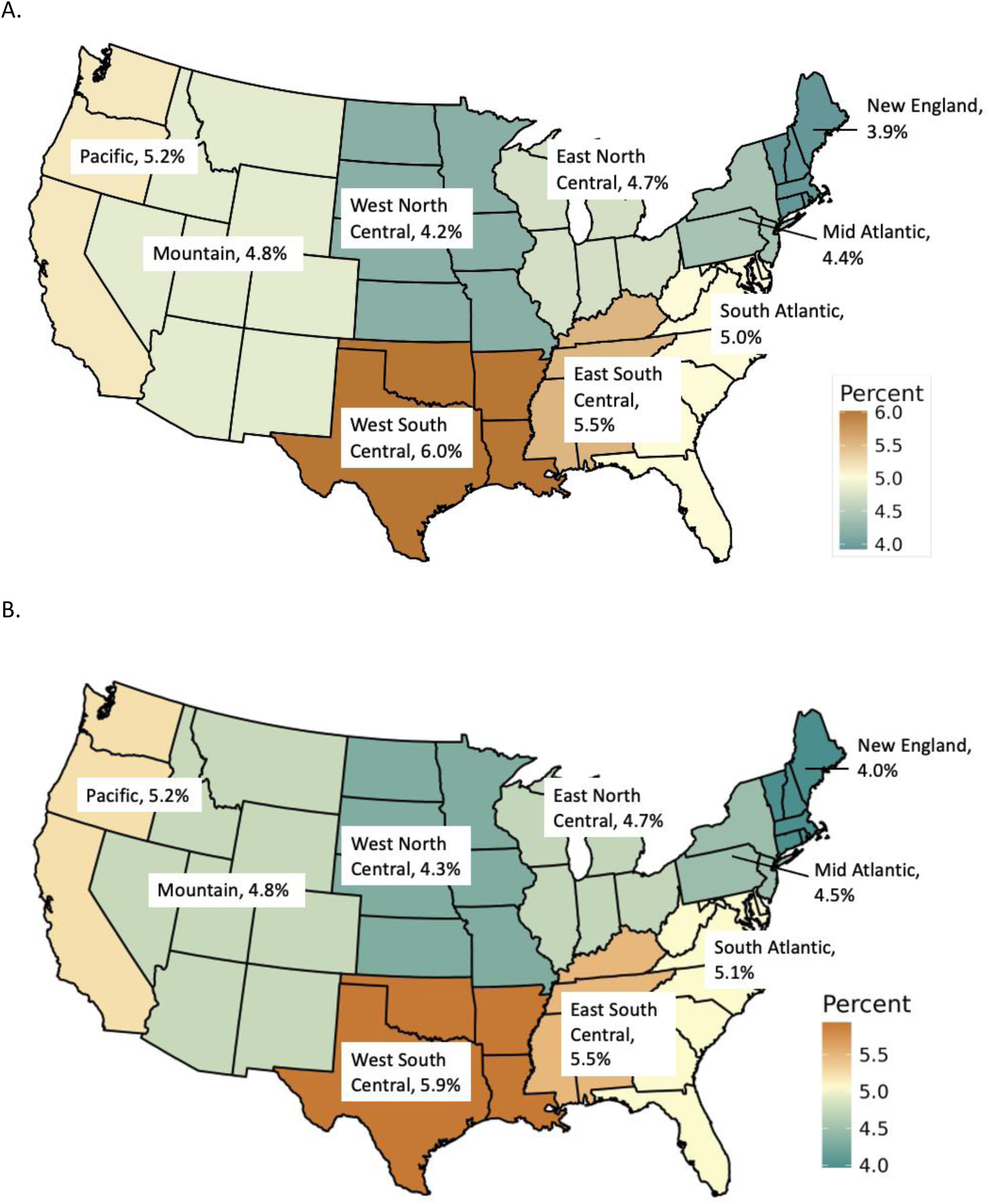
Percentage of Incident Gabapentinoid Initiators within 180 days after Stroke Discharge by U.S. Census Division A) Crude Percentage B) Adjusted Percentage standardized by Age, US Division, mRS, and Discharge Destination using East North Central covariate distribution

## DISCUSSION

In a national U.S. Medicare sample of stroke survivors 65 years of age and older, the percentage who initiated gabapentinoids within six months of discharge increased from 3.6% in 2009 to 5.8% in 2022. The highest initiation percent was observed in 2020. Geographic variation was also evident with higher initiations in the West South Central U.S. region.

The increasing percentage of gabapentinoid initiation observed in this population is concerning, given the limited evidence base to support such a high rate of use. Prophylactic use of anti-seizure medications post-stroke is not recommended.^31^ For patients with recurrent seizures, medication choice should be based on individual characteristics. Gabapentinoids in this setting seem to be primarily used for the management of pain, rather than seizure control.

The Guidelines for Adult Stroke Rehabilitation and Recovery recommend these drugs as a second-line treatment for central post-stroke pain (CPSP), alongside carbamazepine and phenytoin.^32^ However, evidence supporting the efficacy of proposed treatments is limited, and serious side effects have been observed, such as atrial fibrillation among older adults with no pre-existing cardiovascular condition who are new initiators.^33^ Gabapentin specifically has not been adequately studied for CPSP.^32^ Although pregabalin is FDA-approved for certain types of neuropathic pain,^8^ its effectiveness in CPSP is uncertain, and its main benefit seems to derive from improved sleep and anxiety.^34^

Consistent with findings from previous studies, we observed that gabapentin was prescribed far more often than pregabalin.^16,35^ It is estimated that 98.3% of gabapentin prescriptions are for off-label use in chronic pain, including conditions for which pregabalin is formally indicated (fibromyalgia, diabetic neuropathy).^16^ This pattern is likely influenced by multiple factors, including the federal classification of pregabalin as a Schedule V controlled substance, as well as substantial cost differences between the drugs. While gabapentin is widely available in generic form, pregabalin remains available only as a brand-name drug (Lyrica).^6^

In our study, initiation was higher among patients with baseline comorbidities for which these agents are commonly prescribed (i.e, depression, rheumatoid arthritis/osteoarthritis, and fibromyalgia/chronic pain/fatigue). Interestingly, the percentage of individuals with documented fibromyalgia/chronic pain/fatigue at baseline increased over time, which may explain some of the increase in gabapentinoid use. We also observed higher initiation among patients who were discharged to long-term care and inpatient rehabilitation, potentially reflecting greater stroke severity. The prevalence of post-stroke pain is higher among those with more severe strokes, as well as among those with a prior history of chronic pain.^5^

In addition to comorbidities, we identified significant demographic and geographic variation in initiation percentages. Initiation was higher among females, American Indian or Alaska Natives, and individuals from the West South Central region. These findings are consistent with previous research, which similarly identified higher percentages among women and residents of the Southern US.^18,36^ However, most studies have relied on data from a younger, commercially insured population.^11,17,18^ Evidence on older Medicare beneficiaries is limited by shorter observation periods. Most available research focuses on concomitant use of gabapentinoids and other potentially inappropriate drugs or patients with specific pain-related conditions, like fibromyalgia, neuropathy, and post-operative pain.^9,37,38^

Notably, a large percentage of patients in the sample were already prescribed gabapentinoids before their AIS episode. Although excluded from our primary analysis, this finding highlights the widespread use of these medications in older adults. The safety profile of gabapentinoids is concerning in this population, who are especially vulnerable to chronic comorbidities following stroke.^39^ Gabapentin has been associated with cognitive decline and increased risk of dementia.^40,41^ Additionally, to achieve adequate pain control, gabapentinoids are frequently prescribed with other CNS depressants, such as opioids and benzodiazepines, with 10 to 30% of older adults reporting concomitant use.^42,43^ Such polypharmacy is strongly discouraged by the American Geriatrics Society and the FDA, given the risks of severe respiratory depression and death.^44–46^

Several states have implemented regulatory measures to mitigate increasing misuse in response to these concerns.^47^ Alabama, Kentucky, Michigan, North Dakota, Tennessee, Virginia, and West Virginia have classified gabapentin as a Schedule V controlled substance, and many other states require mandatory prescription drug monitoring program (PDMP) reporting.^48^ These measures appear to influence prescribing behavior in Medicare beneficiaries, with Schedule V classification potentially associated with a greater reduction in mean total days’ supply.^49^ However, some specialists fear that reclassifying gabapentin as a controlled substance at the federal level may yield the opposite effect, further restricting its use and driving reliance on opioids.^50^

Given the largely off-label nature of gabapentinoid prescription, important questions remain regarding the appropriateness of treatment, as well as optimal dosing and duration. Therefore, understanding geographic and temporal variations in prescribing is critical, as it may reflect differences in clinical practice and quality of care, and guide resource allocation and targeted intervention efforts. Prior to our study, there have been no attempts to characterize gabapentinoid initiation among post-stroke patients aged 65 and older. Leveraging Medicare claims data over a decade, we provide valuable insights into real-world prescribing trends in this high-risk population.

### Limitations

Our study has several limitations. First, we analyzed administrative claims data for individuals 65 years and older enrolled in Medicare fee-for-service and prescription drug plans, so this study may not be generalized to other populations, Medicare Advantage enrollees, and those without Medicare prescription drug coverage. Second, we acknowledge the possibility of unmeasured factors that could not be controlled due to data limitations. Medicare administrative claims data is collected for billing purposes; therefore, some clinical data elements may be incomplete or missing. For example, the prescription claims do not contain information on medication indications, making it difficult to identify if the reason for the prescription was related to the AIS event or other concomitant comorbidities. Additional data elements would help understand gabapentinoid prescription patterns. However, we could still standardize our measurements for relevant factors, including age, geography, disability scores, and discharge destination. Third, not all patients in the analytic sample were followed for 180 days, as death was a competing event. Fourth, the medication claims data used for this study included records of all prescriptions billed through insurance for individuals included in the study, which is a reliable measure of new medication initiation patterns. Medications paid for out-of-pocket by the beneficiaries or provided through inpatient hospitals, Veterans Affairs, or other supplemental insurance would not be included in Medicare Part D data.

## CONCLUSION

Post-stroke gabapentinoid prescriptions have increased over time among older-age U.S. Medicare beneficiaries. We observed variation in initiation by U.S. Division, with the percentage of gabapentinoid initiation being highest in the West South Central and lowest in New England U.S. Census Division. In light of increased gabapentin use over time and variation in practice, future studies should examine the safety and effectiveness of gabapentinoids in older stroke survivors.

## Data Availability

The data supporting this study's findings were collected by the U.S. Centers for Medicare & Medicaid Services (CMS) and were made available by CMS with no direct identifiers. We cannot share the data, which were used under license from CMS for this study.

## ACKNOWLEDGEMENTS

J.D.B. completed the data analysis, drafted, edited, and revised the manuscript for intellectual content. R.B.R. drafted, edited, and revised the manuscript for intellectual content. M.A.D. revised the manuscript for intellectual content. J.P.N. facilitated data access and revised the manuscript for intellectual content. S.H. was involved in study design and conceptualization. A.C.T. was involved in the study’s conceptualization and revision for intellectual content. L.M.V.R.M. was involved in study design and conceptualization, obtaining data access, and supervising the study development and manuscript drafting for intellectual content.

## REFERENCES

1. 2025 Heart Disease and Stroke Statistics: A Report of US and Global Data From the American Heart Association | Circulation. Accessed April 11, 2025. https://www.ahajournals.org/doi/10.1161/CIR.0000000000001303

2. Ali M, Tibble H, Brady MC, et al. Prevalence, Trajectory, and Predictors of Poststroke Pain: Retrospective Analysis of Pooled Clinical Trial Data Set. Stroke. 2023;54(12):3107–3116. doi:10.1161/STROKEAHA.123.043355

3. Indredavik B, Rohweder G, Naalsund E, Lydersen S. Medical Complications in a Comprehensive Stroke Unit and an Early Supported Discharge Service. Stroke. 2008;39(2):414–420. doi:10.1161/STROKEAHA.107.489294

4. Paolucci S, Iosa M, Toni D, et al. Prevalence and Time Course of Post-Stroke Pain: A Multicenter Prospective Hospital-Based Study. Pain Med Malden Mass. 2016;17(5):924–930. doi:10.1093/pm/pnv019

5. Klit H, Finnerup NB, Jensen TS. Central post-stroke pain: clinical characteristics, pathophysiology, and management. Lancet Neurol. 2009;8(9):857–868. doi:10.1016/S1474-4422(09)70176-0

6. A Clinical Overview of Off-label Use of Gabapentinoid Drugs | Less is More | JAMA Internal Medicine | JAMA Network. Accessed March 19, 2025. https://jamanetwork.com/journals/jamainternalmedicine/article-abstract/2728959

7. Approved Labeling: Gabapentin. Food and Drug Administration (FDA). 2011. https://www.accessdata.fda.gov/drugsatfda_docs/label/2011/020235s036,020882s022,021129s022lbl.pdf

8. Full prescribing information: Pregabalin. Food and Drug Administration (FDA). 2012. https://www.accessdata.fda.gov/drugsatfda_docs/label/2012/021446s028lbl.pdf

9. Bongiovanni T, Gan S, Finlayson E, et al. Trends in the Use of Gabapentinoids and Opioids in the Postoperative Period Among Older Adults. JAMA Netw Open. 2023;6(6):e2318626. doi:10.1001/jamanetworkopen.2023.18626

10. Oh Gy, Moga DC, Fardo DW, Harp JP, Abner EL. The Association of Gabapentin Initiation with Cognitive and Behavioral Changes in Older Adults with Cognitive Impairment: A Retrospective Cohort Study. Drugs Aging. 2024;41(7):623–632. doi:10.1007/s40266-024-01130-z

11. Johansen ME, Maust DT. Update to Gabapentinoid Use in the United States, 2002-2021. Ann Fam Med. 2024;22(1):45–49. doi:10.1370/afm.3052

12. Johansen ME. Gabapentinoid Use in the United States 2002 Through 2015. JAMA Intern Med. 2018;178(2):292–294. doi:10.1001/jamainternmed.2017.7856

13. Bates D, Schultheis BC, Hanes MC, et al. A Comprehensive Algorithm for Management of Neuropathic Pain. Pain Med Off J Am Acad Pain Med. 2019;20(Suppl 1):S2–S12. doi:10.1093/pm/pnz075

14. Oral and Topical Treatment of Painful Diabetic Polyneuropathy: Practice Guideline Update Summary | Neurology. Accessed July 22, 2025. https://www-neurology-org.treadwell.idm.oclc.org/doi/10.1212/WNL.0000000000013038

15. Medicine Use and Spending in the U.S. Accessed March 23, 2025. https://www.iqvia.com/insights/the-iqvia-institute/reports-and-publications/reports/medicine-use-and-spending-in-the-us-a-review-of-2018-and-outlook-to-2023

16. Zhou L, Bhattacharjee S, Kwoh CK, et al. Trends, Patient and Prescriber Characteristics in Gabapentinoid Use in a Sample of United States Ambulatory Care Visits from 2003 to 2016. J Clin Med. 2019;9(1):83. doi:10.3390/jcm9010083

17. Zhao D, Baek J, Hume AL, McPhillips EA, Lapane KL. Geographic Variation in the Use of Gabapentinoids and Opioids for Pain in a Commercially Insured Adult Population in the United States. J Pain Res. 2022;15:443–454. doi:10.2147/JPR.S345521

18. Pauly NJ, Delcher C, Slavova S, Lindahl E, Talbert J, Freeman PR. Trends in Gabapentin Prescribing in a Commercially Insured U.S. Adult Population, 2009-2016. J Manag Care Spec Pharm. 2020;26(3):10.18553/jmcp.2020.26.3.246. doi:10.18553/jmcp.2020.26.3.246

19. Von Elm E, Altman DG, Egger M, et al. The Strengthening the Reporting of Observational Studies in Epidemiology (STROBE) Statement: Guidelines for Reporting Observational Studies. Ann Intern Med. 2007;147(8):573. doi:10.7326/0003-4819-147-8-200710160-00010

20. Centers for Medicare and Medicaid Services. Centers for Medicare and Medicaid Services. Accessed January 7, 2025. https://www.cms.gov

21. Medicare Provider Analysis and Review (MedPAR). ResDAC (Research Data Assistance Center). Accessed January 7, 2025. https://resdac.org/cms-data/files/medpar

22. McCormick N, Bhole V, Lacaille D, Avina-Zubieta JA. Validity of Diagnostic Codes for Acute Stroke in Administrative Databases: A Systematic Review. Quinn TJ, ed. PLOS ONE. 2015;10(8):e0135834. doi:10.1371/journal.pone.0135834

23. Moura LMVR, Donahue MA, Yan Z, et al. Comparative Effectiveness and Safety of Seizure Prophylaxis Among Adults After Acute Ischemic Stroke. Stroke. 2023;54(2):527–536. doi:10.1161/STROKEAHA.122.039946

24. Eicheldinger C, Bonito A. More accurate racial and ethnic codes for Medicare administrative data. Health Care Financ Rev. 2008;29(3):27–42.

25. Chronic Conditions Data Warehouse. Chronic Conditions Data Warehouse. July 1, 2025. Accessed January 7, 2025. https://www2.ccwdata.org/web/guest/home/

26. Rankin J. Cerebral Vascular Accidents in Patients over the Age of 60: II. Prognosis. Scott Med J. 1957;2(5):200–215. doi:10.1177/003693305700200504

27. De Haan R, Limburg M, Bossuyt P, Van Der Meulen J, Aaronson N. The Clinical Meaning of Rankin ‘Handicap’ Grades After Stroke. Stroke. 1995;26(11):2027–2030. doi:10.1161/01.STR.26.11.2027

28. Saver JL, Chaisinanunkul N, Campbell BCV, et al. Standardized Nomenclature for Modified Rankin Scale Global Disability Outcomes: Consensus Recommendations From Stroke Therapy Academic Industry Roundtable XI. Stroke. 2021;52(9):3054–3062. doi:10.1161/STROKEAHA.121.034480

29. Habib M, Cazé De Medeiros R, Muhammad Ahsan S, et al. A Claims-Based Machine Learning Classifier of Modified Rankin Scale in Acute Ischemic Stroke. Published online February 11, 2025. doi:10.1101/2025.02.06.25321827

30. Statistical Power Analysis for the Behavioral Sciences - National Institutes of Health. Accessed July 29, 2025. https://onesearch.nihlibrary.ors.nih.gov/discovery/fulldisplay/cdi_askewsholts_vlebooks_9781134742776/01NIH_INST:NIH

31. Powers WJ, Rabinstein AA, Ackerson T, et al. Guidelines for the Early Management of Patients With Acute Ischemic Stroke: 2019 Update to the 2018 Guidelines for the Early Management of Acute Ischemic Stroke: A Guideline for Healthcare Professionals From the American Heart Association/American Stroke Association. Stroke. 2019;50(12):e344–e418. doi:10.1161/STR.0000000000000211

32. Guidelines for Adult Stroke Rehabilitation and Recovery. doi:10.1161/STR.0000000000000098

33. Ortiz de Landaluce L, Carbonell P, Asensio C, Escoda N, López P, Laporte JR. Gabapentin and Pregabalin and Risk of Atrial Fibrillation in the Elderly: A Population-Based Cohort Study in an Electronic Prescription Database. Drug Saf. 2018;41(12):1325–1331. doi:10.1007/s40264-018-0695-6

34. Kim JS, Bashford G, Murphy KT, Martin A, Dror V, Cheung R. Safety and efficacy of pregabalin in patients with central post-stroke pain. Pain. 2011;152(5):1018–1023. doi:10.1016/j.pain.2010.12.023

35. Huang LL, Wright JA, Fischer KM, et al. Gabapentinoid Prescribing Practices at a Large Academic Medical Center. Mayo Clin Proc Innov Qual Outcomes. 2023;7(1):58–68. doi:10.1016/j.mayocpiqo.2022.12.002

36. Ramdin C, Chen E, Nelson LS, Mazer-Amirshahi M. Gabapentinoid prescribing patterns and predictors utilizing neural networks: An analysis across emergency departments Nationwide between 2012 and 2021. Am J Emerg Med. 2024;85:59–64. doi:10.1016/j.ajem.2024.08.033

37. Zhou L, Bhattacharjee S, Kwoh CK, et al. Association Between Dual Trajectories of Opioid and Gabapentinoid Use and Healthcare Expenditures Among US Medicare Beneficiaries. Value Health. 2021;24(2):196–205. doi:10.1016/j.jval.2020.12.001

38. Chen C, Winterstein AG, Lo-Ciganic WH, Tighe PJ, Wei YJJ. Concurrent use of prescription gabapentinoids with opioids and risk for fall-related injury among older US Medicare beneficiaries with chronic noncancer pain: A population-based cohort study. Nguyen C, ed. PLOS Med. 2022;19(3):e1003921. doi:10.1371/journal.pmed.1003921

39. Divani AA, Majidi S, Barrett AM, Noorbaloochi S, Luft AR. Consequences of Stroke in Community-Dwelling Elderly. Stroke. 2011;42(7):1821–1825. doi:10.1161/STROKEAHA.110.607630

40. Eghrari NB, Yazji IH, Yavari B, Acker GMV, Kim CH. Risk of dementia following gabapentin prescription in chronic low back pain patients. Reg Anesth Pain Med. Published online July 8, 2025. doi:10.1136/rapm-2025-106577

41. Oh Gy, Moga DC, Fardo DW, Abner EL. The association of gabapentin initiation and neurocognitive changes in older adults with normal cognition. Front Pharmacol. 2022;13:910719. doi:10.3389/fphar.2022.910719

42. Chen C, Lo-Ciganic WH, Winterstein AG, Tighe P, Wei YJJ. Concurrent Use of Prescription Opioids and Gabapentinoids in Older Adults. Am J Prev Med. 2022;62(4):519–528. doi:10.1016/j.amepre.2021.08.024

43. Oh GY, Moga DC, Abner EL. Gabapentin utilization among older adults with different cognitive statuses enrolled in the National Alzheimer’s Coordinating Center (2006-2019). Br J Clin Pharmacol. 2023;89(1):410–415. doi:10.1111/bcp.15532

44. Research C for DE and. FDA warns about serious breathing problems with seizure and nerve pain medicines gabapentin (Neurontin, Gralise, Horizant) and pregabalin (Lyrica, Lyrica CR). FDA. Published online August 9, 2024. Accessed March 17, 2025. https://www.fda.gov/drugs/fda-drug-safety-podcasts/fda-warns-about-serious-breathing-problems-seizure-and-nerve-pain-medicines-gabapentin-neurontin

45. Gabapentin, opioids, and the risk of opioid-related death: A population-based nested case-control study - PubMed. Accessed July 29, 2025. https://pubmed-ncbi-nlm-nih-gov.treadwell.idm.oclc.org/28972983/

46. American Geriatrics Society 2023 updated AGS Beers Criteria® for potentially inappropriate medication use in older adults. doi:10.1111/jgs.18372

47. McNeilage AG, Sim A, Nielsen S, Murnion B, Ashton-James CE. Experiences of misuse and symptoms of dependence among people who use gabapentinoids: A qualitative systematic review. Int J Drug Policy. 2024;133:104605. doi:10.1016/j.drugpo.2024.104605

48. Peckham AM, Ananickal MJ, Sclar DA. Gabapentin use, abuse, and the US opioid epidemic: the case for reclassification as a controlled substance and the need for pharmacovigilance. Risk Manag Healthc Policy. 2018;11:109–116. doi:10.2147/RMHP.S168504

49. Grauer JS, Cramer JD. Association of State-Imposed Restrictions on Gabapentin with Changes in Prescribing in Medicare. J Gen Intern Med. 2022;37(14):3630–3637. doi:10.1007/s11606-021-07314-2

50. Shaw G. Should Gabapentin Be a Controlled Substance? Neurol Today. 2022;22(8):1. doi:10.1097/01.NT.0000830928.96207.f3

